# Infant Vaccination Does Not Predict Increased Infant Mortality Rate: Correcting Past Misinformation

**DOI:** 10.1101/2021.09.03.21263082

**Authors:** Ella Nysetvold, Tess Mika, Weston Elison, Daniel Garrett, Justin Hunt, Inori Tsuchiya, S William Brugger, Mary F Davis, Samuel H Payne, Elizabeth G Bailey

## Abstract

Despite extensive scientific research supporting the safety and effectiveness of approved vaccines, debates about their use continue in the public sphere. A paper prominently circulated on social media concluded that countries requiring more infant vaccinations have higher infant mortality rates (IMR), which has serious public health implications. However, inappropriate data exclusion and other statistical flaws in that paper merit a closer examination of this correlation. We re-analyzed the original data used in Miller and Goldman’s study to investigate the relationship between vaccine doses and IMR. We show that the sub-sample of 30 countries used in the original paper was an unlikely random sample from the entire dataset, as the correlation coefficient of 0.49 reported in that study would only arise about 1 in 100,000 times from random sampling. If we investigate only countries with high or very high development, human development index explains the variability in IMR, and vaccine dose number does not. Next, we show IMR as a function of countries’ actual vaccination rates, rather than vaccination schedule, and show a strong negative correlation between vaccination rates and IMR. Finally, we analyze United States IMR data as a function of Hepatitis B vaccination rate to show an example of increased vaccination rates corresponding with reduced infant death over time. From our analyses, it is clear that vaccination does not predict higher IMR as previously reported.

## Introduction

### Vaccines as a Critical Public Health Issue

Development of vaccinations are viewed as one of the greatest public health successes of all time. Widespread immunization has resulted in the control of many infectious diseases that were previously devastating and lethal, including smallpox, poliomyelitis, measles, rubella, tetanus, diphtheria, Haemophilus influenzae type b, and others(1–3). However, anti-vaccination movements have existed since vaccines were first introduced, and recent waves of this skepticism have led to the resurgence of diseases that were previously controlled(3,4). Recently, this public debate has intensified due to the rapid development and distribution of the COVID-19 vaccine(5).

The term “vaccine hesitancy” has been used to describe the uneasiness of individuals and parents who are unsure about vaccination(4,6). Understanding the factors that lead to vaccine hesitancy has been difficult and complex, and researchers have discovered there are many context-specific and variable factors at play that impact vaccination decisions and behavior, including understanding of scientifically-based risks versus benefits, perceived personal risks versus benefits, and concerns about the vaccination schedule(4,6). This hesitancy has been seen to affect behavior. For example, Martin and Petrie found that mistrust of vaccine benefits and worries about unforeseen future effects of vaccines were statistically predictive of past vaccine refusal and future intentions to refuse vaccination(7).

In the case of vaccines, there can be much more at stake than just the impact on one individual in a community. Vaccination of a large portion of the population (e.g. > 90%) protects the entire population by eliminating disease transmission; this is essential to help those who are medically unable to be vaccinated(8). This indirect protection and community benefit has been observed with various vaccines(9–12), demonstrating that vaccine refusal affects more than just the individual who does not get immunized. For example, Salmon et al. examined the effect of vaccine exemption on both the individual and their community(13). The authors concluded that those who claimed vaccination exemption status were 35 times more likely to contract measles; however, if the number of individuals claiming exemption were to double, even nonexempt individuals could see up to a 30% increase in the incidence of measles. Overall, disease outbreaks are more likely in areas that contain larger numbers of unvaccinated individuals (e.g., Centers for Disease Control and Prevention: retrieved from https://www.cdc.gov/measles/cases-outbreaks.html). Thus, addressing vaccine hesitancy by increasing public confidence in vaccine safety has the potential to positively impact public health and save lives(14).

### Vaccine Misinformation and its Spread

Exposure to anti-vaccine information can directly affect vaccine intentions(15), and exposure to misinformation is more widespread than ever with increased use of the internet and social media(5,16,17). Not only can any information be shared on social media, regardless of its validity, but information can also be amplified quickly and spread virally(18–20). A 2018 study found that sophisticated bots and content polluters are more likely to post about vaccines than average Twitter users, often with anti-vaccine content(21). Research suggests that automated users are at least partially inflating anti-vaccine content and amplifying misinformation online, and this can have serious public health implications(22). This widely disseminated misinformation makes it difficult for individuals to determine which sources of information to trust and can affect their vaccine decisions(23,24).

### Miller and Goldman’s 2011 Paper and the Purpose of this Study

In their 2011 paper, Miller and Goldman(25) examined the correlation between infant mortality rate (IMR) and infant vaccine scheduling in various countries. They concluded that vaccine schedules with a greater number of vaccine doses for infants are correlated with higher IMR, proposing the potential for synergistic toxicity of vaccines. This is in sharp contrast to the scientific consensus that vaccines are safe and beneficial for infants even when given with other vaccines(26–29). Although the 2011 study was published in a peer-reviewed journal (Human and Experimental Toxicology), a brief reading of the Miller and Goldman manuscript led us to question the methods, results and conclusions. We observed significant deficiency in the statistical methods. Thus, it is troublesome that this manuscript is in the top 5% of all research outputs since its publication, being shared extensively on social media with tens of thousands of likes and re-shares (see https://acs.altmetric.com/details/406556).

To be trustworthy, science must be self-correcting(30). Sometimes these corrections are a refinement of current understanding (e.g. Einstein’s advances in Physics(31)), and sometimes they are a reversal of incorrect conclusions. This continual revision of the scientific record is normal, and an essential part of the scientific enterprise. It is critical that flawed scientific publications are recognized, as these can cause serious harm(32). In the case of vaccinations, faulty research impacts not just an individual who avoids vaccinations, but also the public health and safety of the population as a whole(8). Due to the disproportionate effect Miller and Goldman’s 2011 paper has had on the public conversation about vaccine safety compared to other scientific publications, we repeated their analysis to examine whether their conclusions are justified.

## Methods

All data and scripts used for the calculations and figures in this manuscript are publicly available on GitHub at https://github.com/PayneLab/vaccine_reevaluation.

### Data Sources

IMR and immunization schedule data from the sources referenced in the original paper were used for Figure 1. The “CIA Country comparison: infant mortality rate data” (2009) was no longer available on www.cia.gov, the website referenced in the original paper. However, the same dataset was found on http://teacherlink.ed.usu.edu/tlresources/reference/factbook/rankorder/2091rank.html. We determined that this was an identical dataset to what was used by Miller and Goldman by manually confirming that the metrics for each of the 30 included countries were identical. For long term preservation, this file has been added to our GitHub repository, see ∼/data/2009_IMR_data.txt.

**Figure 1:**
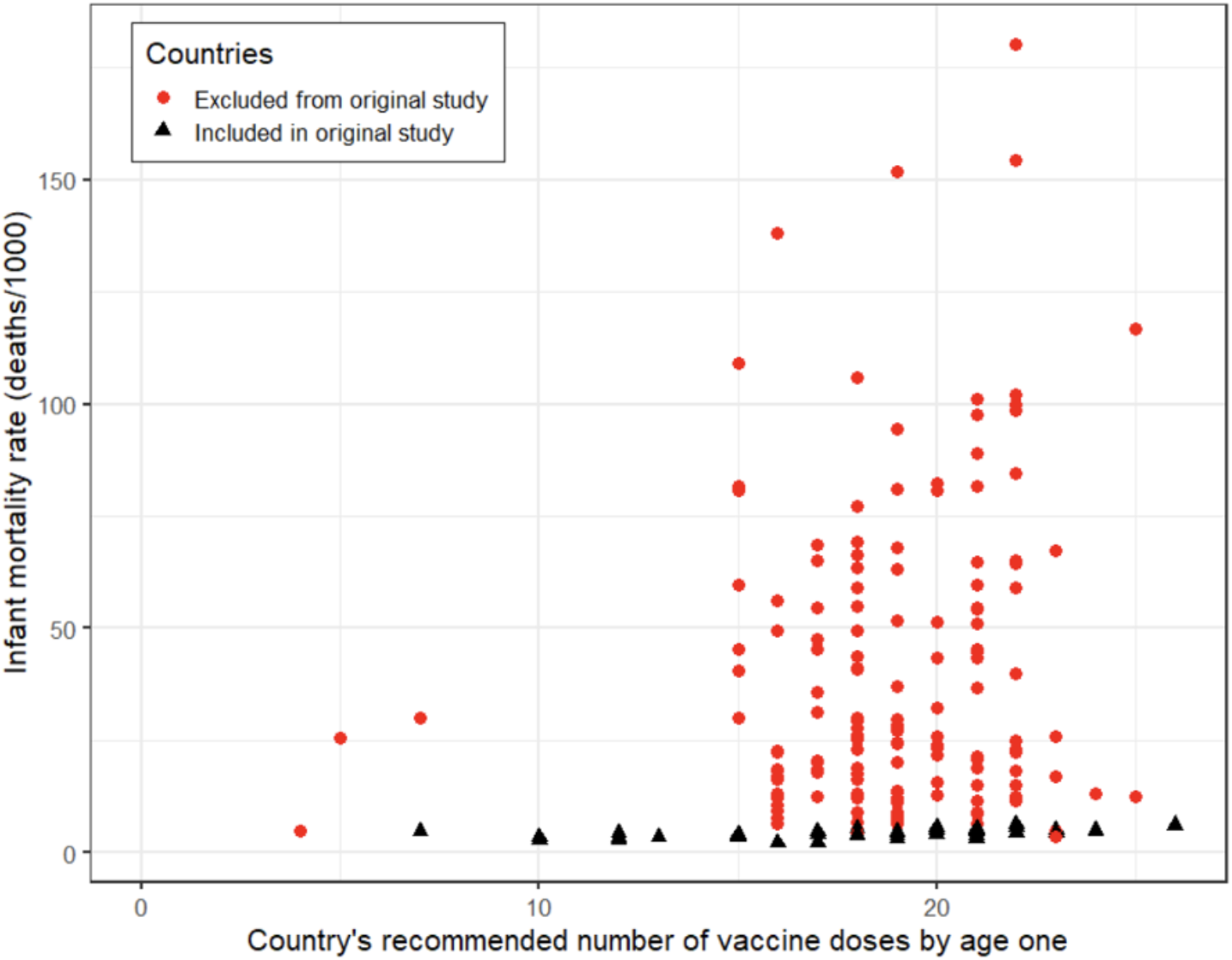
2009 Vaccine Dose and IMR Data Exclusion. The graph above shows a stark contrast between the data used in Miller and Goldman’s study (in black), and the data in the dataset that was available for that study (in red). Using only a very small subset of the available data, the original study showed a moderate correlation between the number of scheduled vaccine doses and IMR (R^2^ = 0.493). However, when the full dataset is included, the correlation is near zero (R^2^ = 0.026), indicating that these two variables are unrelated. Thus, the exclusion of data dramatically changed the conclusion of the data analysis.

Immunization schedule data, detailing the ages at which each vaccine is recommended within each country, was collected from the “WHO/UNICEF Immunization Summary: A Statistical Reference Containing Data Through 2008 (The 2010 edition),” as referenced in the original paper (https://data.unicef.org/wp-content/uploads/2015/12/Immunization_Summary_2008_53.pdf). This file is now saved in our GitHub, see ∼/data/Immunization_Summary_2008_53.pdf.

The covariates used in the multiple linear regression analyses of Tables 1 and 2 were found using the following sources. Human development index (HDI) values were obtained from the United Nation’s 2009 Human Development Report (33). Gini index values (represented income inequality) were downloaded from the World Bank data repository: https://data.worldbank.org/indicator/SI.POV.GINI. We used Gini index values from 2009 or, if unavailable, the most recent value before 2009. Heathcare Access and Quality index (HAQ) values from 2010 were downloaded from Our World in Data: https://ourworldindata.org/grapher/healthcare-access-and-quality-index.

**Table 1:** Multiple linear regression predicting infant mortality rate with vaccine schedule and human development.

| $R^2$ | Adj $R^2$ | Predictor | Coefficient | Std. Error | t value | p value |
| --- | --- | --- | --- | --- | --- | --- |
| 0.58 | 0.57 | (Intercept) | 89.06 | 10.04 | 8.87 | 2.60E-13 |
|  |  | HDI | -89.43 | 9.45 | -9.47 | 1.94E-14 |
|  |  | # Vaccine Doses | 0.02 | 0.14 | 0.13 | 0.90 |

*df = 75 (5 countries excluded due to missing data)*

**Table 2:** Multiple linear regression predicting infant mortality rate with vaccine schedule, human development, income inequality, and healthcare access and quality.

| $R^2$ | Adj $R^2$ | Predictor | Coefficient | Std. Error | t value | p value |
| --- | --- | --- | --- | --- | --- | --- |
| 0.69 | 0.67 | (Intercept) | 73.95 | 15.96 | 4.63 | 2.60E-5 |
|  |  | HDI | -52.70 | 32.65 | -1.61 | 0.11 |
|  |  | Gini Index | 0.10 | 0.10 | 0.98 | 0.33 |
|  |  | HAQ Index | -0.24 | 0.21 | -1.17 | 0.25 |
|  |  | # Vaccine Doses | -0.12 | 0.11 | -1.10 | 0.28 |

*df = 50 (28 countries excluded due to missing data)*

Vaccine doses administered, as used in Figure 3, were downloaded from UNICEF data warehouse: https://data.unicef.org/resources/data_explorer/unicef_f/. We selected the following variables for download:

- infant mortality rate,
- under-five mortality rate,
- child mortality rate (aged 1-4 years),
- Percentage of live births who received bacille Calmette-Guerin (vaccine against tuberculosis),
- Percentage of surviving infants who received the first dose of DTP-containing vaccine,
- Percentage of surviving infants who received the third dose of DTP-containing vaccine,
- Percentage of surviving infants who received the third dose of hep B-containing vaccine,
- Percentage of live births who received hepatitis-B-containing vaccine within 24 hours of birth,
- Percentage of surviving infants who received the third dose of Hib-containing vaccine,
- Percentage of surviving infants who received the first dose of inactivated polio-containing vaccine,
- Percentage of surviving infants who received the first dose of measles-containing vaccine,
- Percentage of children who received the 2nd dose of measles-containing vaccine, as per administered in the national schedule,
- Percentage of surviving infants who received the third dose of pneumococcal conjugate-containing vaccine (PCV),
- Percentage of surviving infants who received the third dose of inactivated polio-containing vaccine,
- Percentage of surviving infants who received the first dose of rubella-containing vaccine,
- Percentage of surviving infants who received the last dose of rotavirus-containing vaccine (2nd or 3rd dose depending on vaccine used),
- Percentage of surviving infants who received yellow fever-containing vaccine (for countries at risk and where the vaccine is in the national schedule)

**Figure 2.**
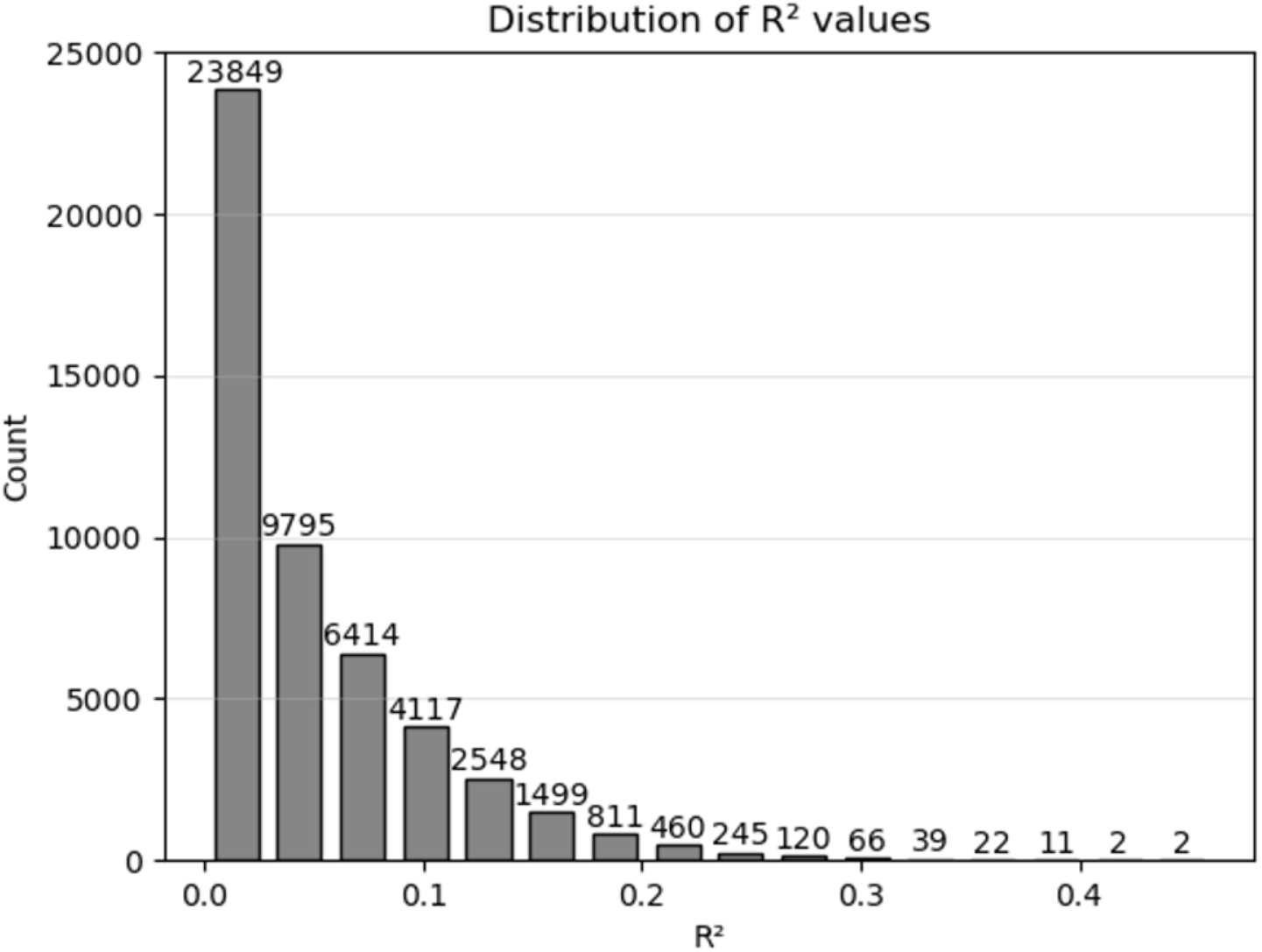
Distribution of R^2^ values from 50,000 random samples. The graph above demonstrates the degree to which R^2^ = 0.493 may be considered an outlier. Of 50,000 random samples of the full dataset, the most extreme R^2^ observed was 0.461, indicating that such a result as reported by Miller and Goldman is statistically improbable.

**Figure 3.**
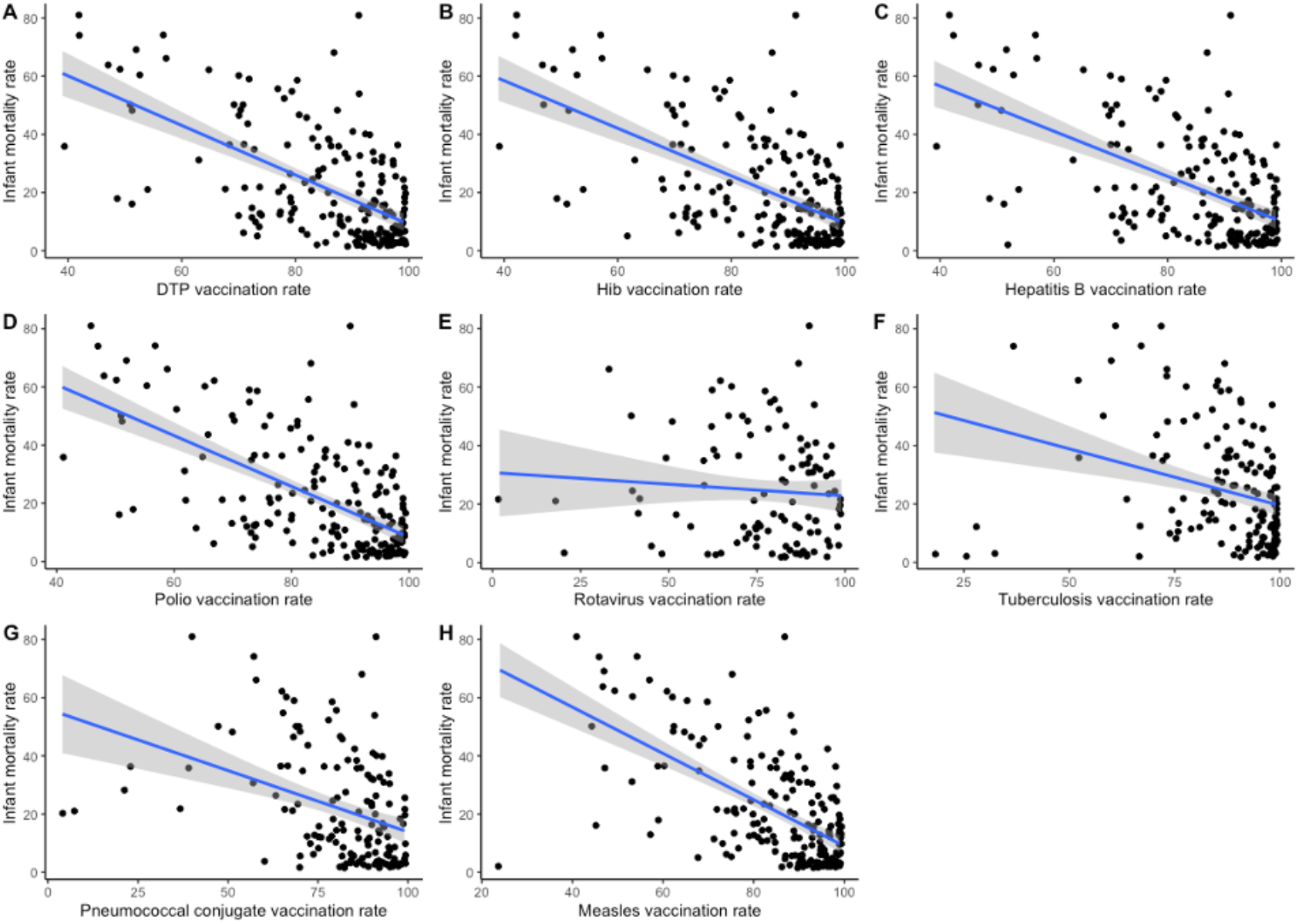
Vaccination rate and infant mortality. For 8 vaccines with global vaccination rate data, we plotted the association with each nation’s infant mortality rate. We also calculated the linear regression statistic for each data set: DPT vaccine correlation coefficient = -0.85 and p = 2.58E-16, Hib vaccine correlation coefficient = -0.82 and p = 3.06E-15, HepB vaccine correlation coefficient = -0.78 and p = 1.472E-12, Polio vaccine correlation coefficient = -0.87 and p < 2.20E-16, Rotavirus vaccine correlation coefficient = -0.079 and p = 0.41, Tuberculosis vaccine correlation coefficient = -0.39 and p = 0.027, PCV vaccine correlation coefficient = -0.42 and p = 0.00029, Measles vaccine correlation coefficient = -0.80 and p = 3.80E-9.

The resulting data have been added to our GitHub repository as

∼/data/Unicef_vaccination_doses_2019.txt

Hepatitis B vaccination rates and longitudinal IMR were retrieved from https://apps.who.int/gho/data/node.main.A828?lang=en and https://childmortality.org respectively. These data have been saved in our GitHub repository, see ∼/data/HepBdata.xls and ∼/data/UNIGME-2020-Country-Sex-specific_U5MR-CMR-and-IMR.xlsx.

### Collecting Vaccine Schedule Data

Combined vaccine dose counts were counted as the number of individual vaccines administered in the combined vaccine (Ex: DTaP = 3 vaccines) multiplied by the number of times the vaccine was scheduled for administration before 12 months of age (Ex: 3 doses of DTaP = 3 doses * 3 vaccines/dose = 9 vaccine doses), as described in Miller and Goldman’s paper. For consistency we used the following criteria for counting vaccine doses, as it matched closest with the numbers included in Miller and Goldman’s paper: only vaccinations scheduled for less than 12 months, or ranges up to 12 months, were included; vaccinations scheduled for high-risk groups, subnational, military groups, travelers, children of carriers, pertussis contraindication, and HIV+ infants were not included. For example, Pneumo_ps is recommended for only high-risk groups in many countries, and so this was not counted in our metric. Doses were manually counted following these criteria and appended to the IMR data and stored as a file called Figure_1_Data.csv.

### Analysis

All of the software and files used in this manuscript, including the code for generating images, is saved in our public GitHub repository https://github.com/PayneLab/vaccine_reevaluation. Data and code used to create Figure 1 and the associated correlation metrics can be found in ∼/code/Make_Figure_1.R. Data and code used to create Figure 2 can be found in ∼/code/Make_Figure_2.R. Sampling of the 30 countries was done at random, and repeated 50,000 times to generate a distribution of potential correlation values. We calculated the simple mean, median, standard deviation, IQR and z-score using the base R functions (see the code). For the vaccination rate and IMR analyses accompanying Figure 3, we calculated simple linear regression with the data downloaded from UNICEF as noted above. All implementation details are available in our GitHub in the script ∼/code/Make_Figure_3.R. We performed multiple linear regression for Tables 1 and 2 using the covariates described above. All code can be found in ∼/code/MLR.R. We calculated the effect of Hepatitis B vaccination rate with a Spearman correlation, using the data from sources above. Data and code used to create Figure 4 can be found in ∼/code/Make_Figure_4.R. IMR rates were separated by sex because Hepatitis B is more prevalent in males(34,35) and the IMR is characteristically distinct by sex.

**Figure 4:**
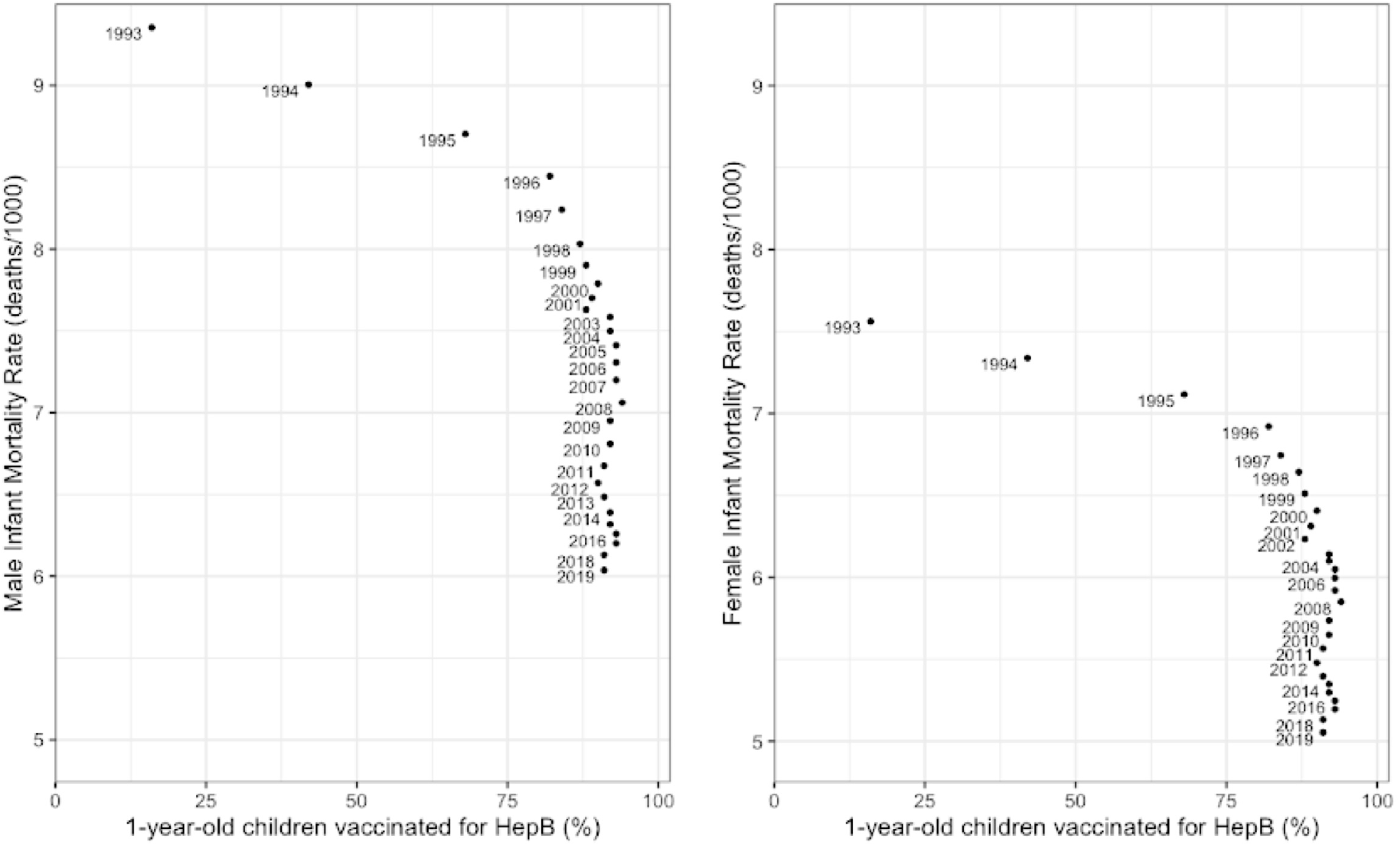
Hepatitis B Vaccine Administration Compared to Male and Female IMR in the United States by Year. These graphs demonstrate a moderate negative correlation between percent of children vaccinated for Hepatitis B and IMR for both males and females in the United States (Spearman Correlation Coefficient = -.665, p-value = .000152).

We checked the assumptions of linear regression for each regression analysis (results in each respective figure or table folder, see also README.md file in github repository for details). For all analyses except rotavirus in Figure 3, the assumption of homoscedasticity was violated. Thus, we used the vcovHC function from the *sandwich* package in R to generate heteroscedasticity robust standard errors for our hypothesis testing (36). Otherwise, all assumptions of linear regression were met.

## Results

A prime conclusion for the manuscript by Miller and Goldman(25) is that “nations that require more vaccine doses tend to have higher infant mortality rates.” At the time of publication, a corrigendum was published to notify readers of unreported affiliations and potential conflicts of interest for the authors(37). However, as we show herein, the most important problem with the manuscript is that their conclusion could only be reached by omitting >80% of the available data. A re-analysis of the full dataset does not support the original conclusion. A re-analysis of only highly or very highly developed countries similarly shows that human development index (HDI) explains the variability in IMR, and more recommended vaccine doses does not predict more infant death.

### Limitations of the Miller and Goldman Study

One of the major errors of Miller and Goldman’s analysis was unexplained data exclusion(25). In their paper, data from only 30 nations was used, despite the fact that data for 185 countries were available in their original data source (Figure 1). Within the text they state that they included “the immunization schedules for the United States and all 33 nations with better IMRs than the United States.” However, there is no scientific reason given for the exclusion of nations with IMR higher than the United States. In fact, the vast majority of the data excluded from analysis had both fewer vaccinations than the US and also a significantly higher IMR than the US. Strikingly, the manuscript itself discusses IMR data for Gambia and Mongolia, both of which were excluded from the statistical analysis, demonstrating that the authors were aware of these data. Excluding data inappropriately can lead to selection bias, contributing to misinformation in the scientific community(36).

### Reanalysis Including Data for Previously Excluded Countries and Relevant Covariates

To re-evaluate the hypothesis that vaccines are associated with infant mortality, we repeated the linear regression analysis using vaccine schedule and IMR data for all countries. This is possible because the original data used by Miller and Goldman is publicly available (see Methods). When all of the data were included, the positive correlation between IMR and immunization schedules disappeared (R^2^ = 0.026 vs. R^2^ = 0.493). While the positive correlation was still significant (p = 0.02), the recommended number of vaccine doses explained less than 3% of the variance. This indicates that there is virtually no relationship between increasing vaccination schedules and infant mortality.

In order to better visualize how extreme Miller and Goldman’s result was even within their own dataset, we randomly sampled 30 countries from the full dataset of 185 countries and computed the linear regression. This sampling was done 50,000 times, and the distribution of regression results was plotted (Figure 2). We then determined the degree to which Miller and Goldman’s result (R^2^ = 0.493) may be considered an outlier. Within this distribution of random samples, the mean R^2^ was 0.049 with a standard deviation of 0.053. We calculated the z-score of 0.493 against our distribution to be 8.3, meaning there is approximately a 1 in a 100,000 chance that this result was achieved with a random sample of the dataset. To verify this, we performed 1 million random samplings, and the most extreme R^2^ observed was 0.577, with only 10 samples’ R^2^ exceeding 0.493. Therefore, we conclude that the sample of 30 countries from the Miller and Goldman analysis is not representative of the full dataset.

It can be appropriate to select a non-random subset of a full dataset with theoretical justification. In their introduction, the original authors discuss that a country’s developmental status has a huge impact on IMR, so it is possible they aimed to investigate the impact of vaccination on IMR in developed countries specifically. However, they did not use a clear inclusion criterion based on development and instead only included countries with lower IMR than the United States. Their initial subset of 34 countries did not include all countries that were rated as *very* highly developed in 2009, and they included Cuba, which was rated as only highly developed (not *very* highly developed as were the others on the list). Thus, to investigate whether the vaccination schedule predicts IMR in developed countries, we ran a linear regression analysis to predict IMR using all 83 countries rated as either highly or *very* highly developed in 2009, with HDI (as an indicator of the degree of development) and number of vaccine doses in the schedule as predictors. As shown in Table 1, this model predicted 57% of the variance in IMR, with development (HDI) significantly contributing to the model and vaccine dosage having no effect.

Infant mortality is a complicated phenomenon influenced by many factors. A literature search revealed that healthcare access and income inequality are two important variables that predict IMR (38–40). Thus, we repeated the multiple linear regression of Table 1 but added the Gini index (to account for income inequality) and the HAQ index (to account for healthcare access and quality) as predictors. As shown in Table 2, this expanded model explained 67% of the variance in IMR for countries with high and very high development. While none of the predictors were individually significant, the overall model (p=2.56E-12) shows that less development, more income inequality, lower healthcare access and quality, and fewer vaccine doses in the schedule were predictive of higher infant mortality rate.

### Vaccination Rate, Not Just Schedule

In the original analysis by Miller and Goldman, they used the vaccine schedule and not the actual data on vaccine doses administered, claiming vaccination rates were high enough it would not affect the results. However, in countries/locales with poor access to health care, the recommended set of vaccinations might not be available to a significant fraction of the population. Therefore, to more clearly answer the question about whether vaccination is related to infant mortality, we compared the vaccination rate for each country against the infant mortality rate. Data from UNICEF includes 2019 global statistics on vaccination rate and IMR for 8 different types of vaccines (see Methods). In agreement with previous literature demonstrating the benefit of vaccines, we show that higher vaccination rates are correlated with lower infant mortality rates for 7 of the 8 vaccinations tested (Figure 3).

### Vaccine Impact Over Time

It is curious that the original Miller and Goldman study did not examine longitudinal data to evaluate their hypotheses. If vaccines were really affecting infant mortality, then the introduction of new vaccines should be correlated with a rise in infant mortality. Therefore, we propose a different test to evaluate the impact of increasing the number of vaccinations. Specifically, we want to evaluate the infant mortality over a time period when a new vaccination becomes common and the impact of that specific addition can be assessed. The Hepatitis B vaccine was introduced in 1981(41) and became common in the United States in the 1990s. We identified a dataset for vaccine doses administered and infant mortality which covers the timeframe of HepB vaccine adoption (see Methods).

We examined the relationship between Hepatitis B vaccination rate (percent of one-year-old children vaccinated) and IMR for male and females in the United States from 1993 - 2019 (Figure 4). For both males and females there is a modest decrease in IMR from 1993 - 1996 as percent of one-year-old children vaccinated for HepB approaches approximately 85%. Between 1996 and 2019, when Hepatitis vaccination remains consistently high, infant mortality drops significantly from 8.5% to 6% in males and from 7% to 5% in females. There are likely confounding factors, and IMR was likely decreasing during this time for other reasons as well. However, these data suggest that if there is any relationship between Hepatitis B vaccination and IMR, it is a lowering of the infant mortality rate. Adding another vaccine to the schedule did not increase infant mortality, which contradicts the original authors’ conclusions about synergistic toxicity. Similar conclusions have been drawn by other studies analyzing vaccine effectiveness(42,43).

## Discussion

Our findings indicate that the conclusions previously suggested by Miller and Goldman(25) are unsupported. More recommended vaccine doses are not associated with an increase in infant mortality. Their conclusion could only be reached by omission of available data (see Figures 1 and 2). When we repeated the analysis with previously excluded data, the positive correlation between vaccination scheduling and IMR disappeared. Even when we focuse on developed countries, vaccine doses are still not predictive of higher IMR, and variance in infant mortality is better explained by country development index (see Tables 1 and 2). We also note that there was a published corrigendum(37) detailing the authors’ affiliations, conflicts of interest, and funding.

While Miller and Goldman claimed that using vaccination rates rather than the schedule would be unlikely to alter their results, we found that using vaccination rates did challenge their 2011 conclusions. When we examined the association between actual vaccination rates and IMR, we found a consistent and strong negative correlation, with higher vaccination rates predicting less infant death (Figure 3). This result better aligns with the scientific consensus about the benefits of vaccination, even with many vaccines given together (26–29). As seen in Figure 3, there is still a lot of variance in IMR unexplained by vaccination rates, so IMR is a complex phenomenon impacted by many country characteristics.

Finally, we presented a case study with longitudinal data demonstrating a lowering of infant mortality in the United States coincident with the widespread adoption of the Hepatitis B vaccine over time (Figure 4). If synergistic toxicity truly exists, as proposed by Miller and Goldman, adding new vaccines to the schedule would have had the opposite effect over time.

The Miller and Goldman study had other limitations not directly addressed in our study. For example, it only looks at the initial effect of vaccines on infants. Vaccines are developed for diseases that affect the entire age spectrum of the population, e.g. infants, adolescents and adults. Therefore, to correctly evaluate the public health impact of vaccination, one would need to include the lives saved when vaccinated individuals no longer acquire these diseases. Such analyses are part of current published literature, e.g. measles cases in the US which dropped dramatically in the 1960s coincident with the introduction of a measles vaccine(44). Another major flaw of the paper’s analysis was simplistic implementation of statistical methods. When studying a question as complicated as infant mortality, including only one variable (vaccination schedule) in regression analyses is not considered best practice. Furthermore, linear regression and correlation coefficients are heavily influenced by outliers(45), which were removed as noted above (see Figure 1). The Miller and Goldman manuscript mentions potential covariates and confounding factors, including many socio-economic factors known to play a critical role in infant mortality(46–49). Unfortunately, even the ones available in their dataset were not included in their analysis. When we include human development index, income inequality, and healthcare access as predictors, vaccine doses either have no impact on IMR, or if anything, correlate with lower IMR (see Tables 1 and 2).

In the context of the current vaccine debate, it is important that accurate information about vaccine safety is accessible. A vast literature exists for the development and clinical testing of individual vaccines (e.g. refs(50–53)), evaluation of vaccination schedules (e.g. refs(26,54–58)) and public health studies testing their efficacy within society (e.g. refs(59–63)). Unfortunately, many individuals get their information from social media, which is not a curated or validated source, and many of these social media users lack the scientific training to evaluate the validity of what they see. In this setting, a single manuscript can have an inordinate impact on public discourse. While corrections and retractions are not always successful at preventing the original misinformation from impacting public debate, repeated corrections and retractions can help alleviate the effects of misinformation(64).

## Data Availability

All data and analyses are availability on GitHub at https://github.com/PayneLab/vaccine_reevaluation.

## Acknowledgements

This work was performed in the BYU Bioinformatics Capstone course and was not supported by an external funding agency.

## References

1. Centers for Disease Control and Prevention (CDC). Ten great public health achievements--United States, 1900-1999. MMWR Morb Mortal Wkly Rep. 1999 Apr 2;48(12):241–3.

2. Centers for Disease Control and Prevention (CDC). Impact of vaccines universally recommended for children--United States, 1990-1998. MMWR Morb Mortal Wkly Rep. 1999 Apr 2;48(12):243–8.

3. Dubé E, Vivion M, MacDonald NE. Vaccine hesitancy, vaccine refusal and the anti-vaccine movement: influence, impact and implications. Expert Rev Vaccines. 2015 Jan;14(1):99–s117.

4. Larson HJ, Cooper LZ, Eskola J, Katz SL, Ratzan S. Addressing the vaccine confidence gap. Lancet. 2011 Aug 6;378(9790):526–35.

5. Puri N, Coomes EA, Haghbayan H, Gunaratne K. Social media and vaccine hesitancy: new updates for the era of COVID-19 and globalized infectious diseases. Hum Vaccin Immunother. 2020 Nov 1;16(11):2586–93.

6. MacDonald NE, SAGE Working Group on Vaccine Hesitancy. Vaccine hesitancy: Definition, scope and determinants. Vaccine. 2015 Aug 14;33(34):4161–4.

7. Martin LR, Petrie KJ. Understanding the Dimensions of Anti-Vaccination Attitudes: the Vaccination Attitudes Examination (VAX) Scale. Ann Behav Med. 2017 Oct;51(5):652–60.

8. Fine P, Eames K, Heymann DL. “Herd immunity”: a rough guide. Clin Infect Dis. 2011 Apr 1;52(7):911–6.

9. Waight PA, Andrews NJ, Ladhani SN, Sheppard CL, Slack MPE, Miller E. Effect of the 13-valent pneumococcal conjugate vaccine on invasive pneumococcal disease in England and Wales 4 years after its introduction: an observational cohort study. Lancet Infect Dis. 2015 May;15(5):535–43.

10. Ali M, Emch M, von Seidlein L, Yunus M, Sack DA, Rao M, et al. Herd immunity conferred by killed oral cholera vaccines in Bangladesh: a reanalysis. Lancet. 2005 Jul 2;366(9479):44–9.

11. Piedra PA, Gaglani MJ, Kozinetz CA, Herschler G, Riggs M, Griffith M, et al. Herd immunity in adults against influenza-related illnesses with use of the trivalent-live attenuated influenza vaccine (CAIV-T) in children. Vaccine. 2005 Feb 18;23(13):1540–8.

12. Lopman BA, Curns AT, Yen C, Parashar UD. Infant rotavirus vaccination may provide indirect protection to older children and adults in the United States. J Infect Dis. 2011 Oct 1;204(7):980–6.

13. Salmon DA, Haber M, Gangarosa EJ, Phillips L, Smith NJ, Chen RT. Health consequences of religious and philosophical exemptions from immunization laws: individual and societal risk of measles. JAMA. 1999 Jul 7;282(1):47–53.

14. Salmon DA, Dudley MZ, Glanz JM, Omer SB. Vaccine Hesitancy: Causes, Consequences, and a Call to Action. Am J Prev Med. 2015 Dec;49(6 Suppl 4):S391–398.

15. Jolley D, Douglas KM. The effects of anti-vaccine conspiracy theories on vaccination intentions. PLoS One. 2014;9(2):e89177.

16. Guess AM, Nyhan B, O’Keeffe Z, Reifler J. The sources and correlates of exposure to vaccine-related (mis)information online. Vaccine. 2020 Nov;38(49):7799–805.

17. Betsch C, Brewer NT, Brocard P, Davies P, Gaissmaier W, Haase N, et al. Opportunities and challenges of Web 2.0 for vaccination decisions. Vaccine. 2012 May 28;30(25):3727–33.

18. Kouzy R, Abi Jaoude J, Kraitem A, El Alam MB, Karam B, Adib E, et al. Coronavirus Goes Viral: Quantifying the COVID-19 Misinformation Epidemic on Twitter. Cureus. 2020 Mar 13;12(3):e7255.

19. Shao C, Ciampaglia GL, Varol O, Yang KC, Flammini A, Menczer F. The spread of low-credibility content by social bots. Nat Commun. 2018 Nov 20;9(1):4787.

20. Beletsky L, Seymour S, Kang S, Siegel Z, Sinha MS, Marino R, et al. Fentanyl panic goes viral: The spread of misinformation about overdose risk from casual contact with fentanyl in mainstream and social media. Int J Drug Policy. 2020 Sep 16;86:102951.

21. Broniatowski DA, Jamison AM, Qi S, AlKulaib L, Chen T, Benton A, et al. Weaponized Health Communication: Twitter Bots and Russian Trolls Amplify the Vaccine Debate. Am J Public Health. 2018 Oct;108(10):1378–84.

22. Jamison AM, Broniatowski DA, Quinn SC. Malicious Actors on Twitter: A Guide for Public Health Researchers. Am J Public Health. 2019 May;109(5):688–92.

23. Kolodziejski L. Beyond the “Hullabaloo” of the Vaccine “Debate”: Understanding Parents’ Assessment of Risks When Making Vaccine Decisions. rhm. 2020 Mar 23;3(1):63–92.

24. Ashfield S, Donelle L. Parental Online Information Access and Childhood Vaccination Decisions in North America: Scoping Review. J Med Internet Res. 2020 Oct 13;22(10):e20002.

25. Miller NZ, Goldman GS. Infant mortality rates regressed against number of vaccine doses routinely given: Is there a biochemical or synergistic toxicity? Hum Exp Toxicol. 2011 Sep;30(9):1420–8.

26. Maglione MA, Das L, Raaen L, Smith A, Chari R, Newberry S, et al. Safety of vaccines used for routine immunization of U.S. children: a systematic review. Pediatrics. 2014 Aug;134(2):325–37.

27. Zhou F, Shefer A, Wenger J, Messonnier M, Wang LY, Lopez A, et al. Economic evaluation of the routine childhood immunization program in the United States, 2009. Pediatrics. 2014 Apr;133(4):577–85.

28. Shinefield H, Black S, Thear M, Coury D, Reisinger K, Rothstein E, et al. Safety and immunogenicity of a measles, mumps, rubella and varicella vaccine given with combined Haemophilus influenzae type b conjugate/hepatitis B vaccines and combined diphtheria-tetanus-acellular pertussis vaccines. Pediatr Infect Dis J. 2006 Apr;25(4):287–92.

29. Becker-Dreps S, Amaya E, Liu L, Moreno G, Rocha J, Briceño R, et al. Changes in childhood pneumonia and infant mortality rates following introduction of the 13-valent pneumococcal conjugate vaccine in Nicaragua. Pediatr Infect Dis J. 2014 Jun;33(6):637–42.

30. Alberts B, Cicerone RJ, Fienberg SE, Kamb A, McNutt M, Nerem RM, et al. Self-correction in science at work. Science. 2015 Jun 26;348(6242):1420–2.

31. Einstein A, Podolsky B, Rosen N. Can Quantum-Mechanical Description of Physical Reality Be Considered Complete? Phys Rev. 1935 May 15;47(10):777–80.

32. Steen RG. Retractions in the medical literature: how many patients are put at risk by flawed research? J Med Ethics. 2011 Nov;37(11):688–92.

33. UNDP, editor. Overcoming barriers: human mobility and development. Houndmills: Palgrave Macmillan; 2009. 217 p. (Human development report).

34. Baig S. Gender disparity in infections of Hepatitis B virus. J Coll Physicians Surg Pak. 2009 Sep;19(9):598–600.

35. Ruggieri A, Gagliardi MC, Anticoli S. Sex-Dependent Outcome of Hepatitis B and C Viruses Infections: Synergy of Sex Hormones and Immune Responses? Front Immunol. 2018 Oct 8;9:2302.

36. Kleiber C, Zeileis A. Applied Econometrics with R. New York: Springer; 2008. 221 p.

37. Corrigendum. Hum Exp Toxicol. 2011 Sep;30(9):1429–1429.

38. Gruber J, Hendren N, Townsend RM. The Great Equalizer: Health Care Access and Infant Mortality in Thailand. Am Econ J Appl Econ. 2014 Jan 1;6(1):91–107.

39. Owusu PA, Sarkodie SA, Pedersen PA. Relationship between mortality and health care expenditure: Sustainable assessment of health care system. PLoS One. 2021;16(2):e0247413.

40. Agustina R, Dartanto T, Sitompul R, Susiloretni KA, Suparmi null, Achadi EL, et al. Universal health coverage in Indonesia: concept, progress, and challenges. Lancet. 2019 Jan 5;393(10166):75–102.

41. Szmuness W, Stevens CE, Zang EA, Harley EJ, Kellner A. A controlled clinical trial of the efficacy of the hepatitis B vaccine (heptavax B): A final report. Hepatology. 1981 Sep;1(5):377–85.

42. Delany I, Rappuoli R, De Gregorio E. Vaccines for the 21st century. EMBO Mol Med. 2014 Jun;6(6):708–20.

43. Epoke J, Eko F, Mboto CI. Vaccinated versus unvaccinated children: how they fare in first five years of life. Trop Geogr Med. 1990 Apr;42(2):182–4.

44. Orenstein WA, Hinman AR, Papania MJ, editors. Evolution of Measles Elimination Strategies in the United States. The Journal of Infectious Diseases. 2004 May 1;189(Supplement_1):S17–22.

45. Altman N, Krzywinski M. Association, correlation and causation. Nat Methods. 2015 Oct;12(10):899–900.

46. Ely DM, Driscoll AK, Matthews TJ. Infant Mortality Rates in Rural and Urban Areas in the United States, 2014. NCHS Data Brief. 2017 Sep;(285):1–8.

47. Overpeck MD, Hoffman HJ, Prager K. The lowest birth-weight infants and the US infant mortality rate: NCHS 1983 linked birth/infant death data. Am J Public Health. 1992 Mar;82(3):441–4.

48. McElroy JA, Bloom T, Moore K, Geden B, Everett K, Bullock LF. Perinatal mortality and adverse pregnancy outcomes in a low-income rural population of women who smoke. Birth Defects Research Part A: Clinical and Molecular Teratology. 2012 Apr;94(4):223–9.

49. He X, Akil L, Aker W, Hwang HM, Ahmad H. Trends in Infant Mortality in United States: A Brief Study of the Southeastern States from 2005–2009. IJERPH. 2015 May 6;12(5):4908–20.

50. Slaoui M, Hepburn M. Developing Safe and Effective Covid Vaccines — Operation Warp Speed’s Strategy and Approach. N Engl J Med. 2020 Oct 29;383(18):1701–3.

51. Skowronski DM, De Serres G. Safety and Efficacy of the BNT162b2 mRNA Covid-19 Vaccine. N Engl J Med. 2021 Apr 22;384(16):1576–7.

52. Szmuness W, Stevens CE, Harley EJ, Zang EA, Oleszko WR, William DC, et al. Hepatitis B vaccine: demonstration of efficacy in a controlled clinical trial in a high-risk population in the United States. N Engl J Med. 1980 Oct 9;303(15):833–41.

53. Paavonen J, Jenkins D, Bosch FX, Naud P, Salmerón J, Wheeler CM, et al. Efficacy of a prophylactic adjuvanted bivalent L1 virus-like-particle vaccine against infection with human papillomavirus types 16 and 18 in young women: an interim analysis of a phase III double-blind, randomised controlled trial. Lancet. 2007 Jun 30;369(9580):2161–70.

54. Rupprecht CE, Briggs D, Brown CM, Franka R, Katz SL, Kerr HD, et al. Evidence for a 4-dose vaccine schedule for human rabies post-exposure prophylaxis in previously non-vaccinated individuals. Vaccine. 2009 Nov 27;27(51):7141–8.

55. Bonanni P, Chiamenti G, Conforti G, Maio T, Odone A, Russo R, et al. The 2016 Lifetime Immunization Schedule, approved by the Italian scientific societies: A new paradigm to promote vaccination at all ages. Hum Vaccin Immunother. 2017 Nov 2;13(11):2531–7.

56. Buttery JP, Lambert SB, Grimwood K, Nissen MD, Field EJ, Macartney KK, et al. Reduction in rotavirus-associated acute gastroenteritis following introduction of rotavirus vaccine into Australia’s National Childhood vaccine schedule. Pediatr Infect Dis J. 2011 Jan;30(1 Suppl):S25–29.

57. Englund JA, Walter EB, Fairchok MP, Monto AS, Neuzil KM. A comparison of 2 influenza vaccine schedules in 6-to 23-month-old children. Pediatrics. 2005 Apr;115(4):1039–47.

58. Avdicová M, Prikazský V, Hudecková H, Schuerman L, Willems P. Immunogenicity and reactogenicity of a novel hexavalent DTPa-HBV-IPV/Hib vaccine compared to separate concomitant injections of DTPa-IPV/Hib and HBV vaccines, when administered according to a 3, 5 and 11 month vaccination schedule. Eur J Pediatr. 2002 Nov;161(11):581–7.

59. Gessner BD, Kaslow D, Louis J, Neuzil K, O’Brien KL, Picot V, et al. Estimating the full public health value of vaccination. Vaccine. 2017 Nov;35(46):6255–63.

60. Ruhnke GW, Coca-Perraillon M, Kitch BT, Cutler DM. Marked reduction in 30-day mortality among elderly patients with community-acquired pneumonia. Am J Med. 2011 Feb;124(2):171-178.e1.

61. Anderson EJ, Daugherty MA, Pickering LK, Orenstein WA, Yogev R. Protecting the Community Through Child Vaccination. Clin Infect Dis. 2018 Jul 18;67(3):464–71.

62. Aaby P, Bukh J, Lisse IM, Smits AJ. Measles vaccination and reduction in child mortality: a community study from Guinea-Bissau. J Infect. 1984 Jan;8(1):13–21.

63. Zanettini C, Omar M, Dinalankara W, Imada EL, Colantuoni E, Parmigiani G, et al. Influenza Vaccination and COVID19 Mortality in the USA. medRxiv. 2020 Jun 26;2020.06.24.20129817.

64. Lewandowsky S, Ecker UKH, Seifert CM, Schwarz N, Cook J. Misinformation and Its Correction: Continued Influence and Successful Debiasing. Psychol Sci Public Interest. 2012 Dec;13(3):106–31.

